# THE SILENT STRUGGLE: EXPLORING THE EFFECTS OF COMMUNICATION BREAKDOWNS IN HEALTHCARE DELIVERY IN THE NORTHERN REGION OF GHANA

**DOI:** 10.64898/2026.06.18.26356013

**Authors:** Issah Salifu, Muhammed Abdulai, Naazia Ibrahim

## Abstract

This qualitative study examines the lived experiences of patients and healthcare professionals regarding communication breakdowns in healthcare delivery in the Northern Region of Ghana, using a phenomenological approach. A purposive sample of 20 participants took part in semi-structured interviews, and the data was thematically analyzed to identify key themes and patterns. The findings reveal that linguistic diversity, low health literacy, and hierarchical communication systems are common causes of miscommunication, misinterpretation, misconceptions and misdiagnoses. Due to logistical challenges and fragmented care coordination, delays in treatment and additional complications emerged as systemic issues.

In addition, participants expressed less satisfaction with their experiences receiving healthcare and emphasized how imprecise instructions and a lack of effective provider-patient communication caused poor adherence to treatment protocols. To overcome linguistic and cultural barriers, the study suggests that healthcare professionals receive focused training in intercultural competency and that interpreters with professional training, local language and cultural knowledge should be strategically hired for provider-patients’ interpretations. Enhancing communication, building patient trust, and improving regional healthcare results largely depend on these initiatives.

## Introduction

The significance of effective communication in health care settings cannot be overemphasized, especially in the context of hospitals in the Northern Region of Ghana. Communication is the cornerstone of patient-healthcare provider interactions, influencing not only the quality of care but also patient satisfaction and health outcomes (Huang et al, 2025). Khorram-Manesh et al. (2024) noted that globally, health communication has become an essential area of research and practice, responding to the multifaceted challenges arising from diverse cultural, social and economic contexts. The World Health Organization emphasizes that clear and respectful communication is essential to ensure patient safety and satisfaction, emphasizing its role in achieving high-quality health care (Omaghomi et al., 2024). In Africa, the importance of effective communication is further amplified by unique challenges such as language diversity, cultural differences, and varying levels of health literacy (Ayo-Farai et al., 2023). These barriers can significantly impede the flow of information between patients and healthcare providers, leading to misunderstandings and sub-optimal care. Studies such as those by Aseeri et al. (2024) and Mustafa et al. (2023) have shown that poor communication can lead to misdiagnosis, inadequate treatment adherence, and overall dissatisfaction with healthcare services.

While studies such as Steinhorst et al. (2021), Jiang et al. (2023) and Abdulai et al. (2019) have explored health communication-related issues in various ways, there are still gaps that need to be addressed to improve the delivery of high-quality health care in the Northern Region of Ghana, where referral hospitals serve as the key health care organizational foundations for the northern region. In the Chinese context, Jiang et al. (2023) investigated the impact of language use on healthcare delivery and found that misunderstandings could lead to inappropriate treatments, misdiagnoses, and overall dissatisfaction with care. Their study was, however, limited to the Chinese context. Again, Steinhorst et al. (2021) also examined health communication in Ghana. Although their study was limited to traditional healers and snakebite patients. Abdulai et al. (2019) qualitative study in the northern Ghanaian context was limited in terms of scope as the Sissala West District of the Upper West Region was the research site and again variants of the Sissala dialects were the only variables that were examined in terms of its effects on the delivery of quality health care. Considering this, existing literature on health communication often overlooks the specific context of northern region of Ghana, creating critical empirical and evidence gaps that this study seeks to fill. By exploring the impact of communication barriers on effective health communication and identifying strategies in this unique cultural context to improve dialogic interactions between patients and providers. Also, despite the recognized role of communication in healthcare delivery, limited empirical research has explored how cultural barriers affect quality health service delivery in tertiary hospitals in the Northern Region of Ghana. The significance of this study lies in its application of dialogic engagement framework to examine the influence of culture on effective communication between patients and health care providers in the Northern Region of Ghana. Findings from this study will not only contribute to localized healthcare improvement but also add to broader scholarly discussions on health communication in culturally diverse settings. In addition, the findings will provide practical insights for strengthening the healthcare systems in the Northern Region of Ghana and guide the development of targeted communication strategies to improve effective communication between patients and healthcare providers.

### Rethinking Health Communication: Cultural Dimensions in Delivering Quality Health Care

This section reviews relevant literature aims to conceptualize health communication, the impact of communication barriers on the effectiveness of health messages and theoretical underpinnings of culture and effective health communication.

Health communication is said to be the process of creating, sharing, and interpreting health-related messages. According to Igwe and Obeagu (2024), effective health communication is essential to promote healthy behaviors, prevent disease, and improve health care outcomes. It involves various stakeholders, including health care providers, patients, policymakers, and the media in achieving quality healthcare delivery within the broader tapestry of healthcare services. Also, Natarajan and Parikh (2013) assert that the field of health communication has evolved significantly, especially with the advent of digital media, which has changed the way health information is disseminated and consumed. In addition, recent literature on health communication emphasizes that health communication is not only about conveying information but also about understanding the needs, preferences, and cultural context of the audiences (McKenzie et al., 2022). This perspective is consistent with social cognitive theory (SCT), which argues that behavior change is influenced by personal factors, environmental influences, and the behavior itself (Bandura, 2004). Therefore, effective health communication must take these aspects into account to promote better health outcomes.

Globally, effective health communication is recognized as a fundamental part of delivering quality health care. Wahyudi et al. (2023) and Larson et al. (2019) have all argued that clear communication could significantly improve patient outcomes, satisfaction, and adherence. This implies that communication that is simple, concise, accurate and unambiguous and purpose driven satisfies the needs of both patients and providers leading to positive outcomes. It is, however, significant to note that challenges such as cultural differences, language barriers, and socioeconomic disparities have complicated the global health communication landscape (Bensley and Brookins-Fisher, 2023; Giger and Haddad, 2020).

In the African context, communication barriers are particularly pronounced due to the continent’s linguistic diversity and cultural differences. Research highlights that language difficulties often impede effective interactions between patients and health care providers, leading to misunderstandings that adversely impact treatment outcomes (Ugwu, 2019; Ward et al., 2019). Additionally, cultural beliefs influence patient perceptions of illness and health care practices, with some communities preferring traditional healing methods over modern medical care (Tanaka et al., 2023). These factors contribute to poor health literacy, exacerbating communication challenges in health care settings (de Vries et al., 2019).

In the context of Northern Ghana, the referral Hospitals are important health care facilities for a diverse population and cultures. It is however, significant to note that some barriers to effective health communication exist in the region. As noted, before, a study conducted by Abdulai et al. (2019) shows that the coexistence of diverse languages and dialects complicate interactions between patients and health care providers. Furthermore, cultural factors play an important role in determining patients’ willingness to engage with health services. Moreover, Misinterpretation could lead to incorrect diagnoses and disrupts treatment plans, ultimately leading to patient dissatisfaction and poor health outcomes (Jiang et al., 2023). Therefore, understanding the specific barriers to effective communication at Tamale Teaching Hospital, Yendi Government and Bimbilla Hospital are important for developing effective communication strategies to improve patient-healthcare provider interactions and overall health care quality.

In conclusion, conceptualizing health communication requires understanding the complex nature of the interaction of various stakeholders in various contexts. This is often influenced by multiple barriers that impact on the effectiveness of message delivery, thus ultimately influencing public health outcomes in a positive or negative way.

### Culture and Health: A Constructivist Approach

The connection between **health and culture** is profound and multifaceted. According to Onuoha et al. (2024) Culture shapes how individuals perceive illness, seek treatment, and engage with health systems. This implies that diagnosis through complete treatment plans is deemed comprehensive and successful only when they reflect the cultural requirements of patients. Recent literature emphasizes that health is not just biological-it’s deeply embedded in social and cultural contexts (Oman, 2025). The significance of culture in understanding health communication is crucial and intricate.

Rehfeldt et al. (2021) define culture as a complex system that includes language, religion, cuisine, social habits, music, and art. Lansford (2022) also states that culture includes the behaviors, norms, beliefs, and social practices that characterize a group of people. This implies that all areas of an individual are covered by culture and for communication to take place, individuals must be culturally competent. Individuals are defined by their culture, and according to Phinney et al. (2022), cultural identity refers to the sense of belonging to a particular culture or ethnicity. Culture plays an important role in shaping an individual’s self-perception and worldview. Studies have emphasized the importance of cultural diversity in fostering creativity and innovation in society (Mahama, 2024). Qorib and Afandi (2024) assert that culture provides shared meanings and values that facilitate communication and cooperation among members of a society. In short, culture is a multifaceted construct that shapes human experiences in many different aspects. To understand its complexity, we must appreciate both its stability and its dynamics in the face of internal and external influences. Cultural beliefs and practices largely influence how patients perceive health and illness. Individuals from different cultures may have different attitudes toward seeking medical care, adherence to treatment, and the role of health care providers (Tanaka et al., 2023). For example, some cultures may favor traditional healing methods over modern medicine, which can place unnecessary stress on the patient-provider relationship and lead to ineffective communication (Cutler et al., 2019).

Panjaitan et al. (2023) assert that health communication, which includes information sharing and the encouragement of health-related behaviours, is an essential component of public health. Cultural norms, values, and beliefs are among the many factors that influence it (Higueras-Castillo, et al., 2024). Creating successful health treatments and guaranteeing fair access to medical care depend on an understanding of how culture affects health communication. By identifying important themes, difficulties, and practical implications, this literature review seeks to throw light on the relationship between culture and health communication. Again, people’s views of health and illness are shaped by their culture, which also affects how people interact with healthcare professionals and understand health messaging. Similarly, cultural attitudes could influence health behaviours including following medical advice or taking part in preventative care programs (Kale et al., 2023; Galesic et al., 2020). For example, some cultures may value traditional healing methods more than biomedical ones, which could cause problems between patients and medical professionals (Kleinman & Benson, 2006). Furthermore, communication about health might be greatly impacted by language limitations. Patients who are not proficient in the dominant language may find it more difficult to comprehend medical information and to properly communicate their symptoms or concerns to healthcare practitioners (Ortega et al., 2022). The significance of culturally sensitive communication techniques that take language differences into account is highlighted by this difficulty. The ability of healthcare professionals to comprehend and effectively address patients’ cultural demands is known as cultural competency (Antón-Solanas, et al., 2022). Nwosu et al. (2025) claim that culturally competent care lowers health outcome disparities, increases treatment adherence, and improves patient satisfaction. Fostering successful communication with diverse communities requires healthcare practitioners to receive training in cultural competence. Culturally appropriate interventions have been linked to improved health outcomes, according to research (Griffith et al., 2024). For instance, a study conducted by Griffith et al. (2024) showed that, in contrast to traditional instructional materials, culturally tailored materials improved African American patients’ understanding of diabetes care. These results emphasize how important it is to incorporate cultural factors into health communication plans.

As stated earlier, this study is also situated within the constructivist approach of culture and health. The constructivist theory holds that knowledge is created via social interactions and experiences rather than passively received (Yakubu, 2025). This theory highlights how people actively interpret health information and meaning based on their experiences, beliefs, and social situations (Sykes et al., 2025).

Constructivism is distinguished by several fundamental ideas: one of the constructivist’s theories of culture this study employed is the Andrian Holliday‘s concept of small and large culture and the Onion metaphor of culture (Holliday, 2022). Adrian Holliday’s concept of **small and large culture** aligns well with this study because, the study seeks to examines the lived experiences of patients and healthcare professionals regarding communication breakdowns in healthcare delivery in the Northern Region of Ghana. Considering this, l*arge culture* refers to broad, often stereotypical national or ethnic identities that are typically reified and static (Kariya and Rappleye, 2020). Holliday critiques this view for promoting essentialism and overlooking the complexity of human interaction (Holliday, 2022). In contrast, *small culture* emphasizes dynamic, localized, and emergent cultural practices formed within specific social groups, such as classrooms, workplaces, or communities (Tiwari and Fahrudin, 2024). These are fluid and shaped by interpersonal relationships and shared experiences. In the context of this study, the concept of small culture will be utilized to examine the influence of culture on effective communication between patients and health care providers in the Northern Region of Ghana. To this end, it will cast light on how the cultural beliefs, values and norms of health service providers and patients influence the meanings they attached to the signs and symbols used between healthcare providers and patients.

This shift from static to dynamic cultural models encourages educators, researchers, and global citizens to move beyond simplistic cultural binaries and engage with the rich, evolving tapestry of human experience.

Also, the onion metaphor of culture has been applied in this study to explain the nuanced lived experiences of both patients and providers in the day-to-day interactions. The idea of the onion metaphor was first conceived by eminent anthropologist Edward T. Hall, who is well-known for his contributions to the field of intercultural communication (Nguyen-Phuong-Mai, 2019). In 1976, Hall presented a conceptualization of culture as a multi-layered onion in his groundbreaking book “Beyond Culture.” The author suggests that culture is made up of both visible and invisible layers that affect people’s behavior, viewpoints, and social interactions (Bettache, 2025). According to Fishman’s (2017), the most visible aspects of culture include things like dress, food, and language usage. As the researcher digs deeper into the investigation, they encounter ever deeper layers spanning social norms, values, beliefs, and viewpoints.

Building on the groundbreaking work of Hall, later researchers have developed the onion metaphor to explore various aspects of culture. The concept was further developed by renowned psychologist and cross-cultural studies expert Geert Hofstede in his landmark work “Cultures and Organisations: Software of the Mind” (1991).

This idea offers a paradigm that is very helpful in understanding cultural dynamics and the complex nature of cross-cultural relationships. By investigating the several levels of cultural dynamics, researchers can gain a more profound comprehension of the underlying ideas, beliefs, and customs that shape people’s behaviours and viewpoints (Nguyen-Phuong-Mai, 2017).

All things considered, the application of the onion metaphor in cultural studies provides a useful framework for understanding the complex and multifaceted nature of cultural processes. This emphasizes the idea that culture is more than just outward manifestations; it has deeper aspects that shape people’s behavior and worldview. In the field of healthcare delivery, effective communication between patients and healthcare providers is critical to ensuring that patients receive high-quality care and achieve positive health outcomes. However, cultural differences may make it more difficult to understand and provide healthcare, which could result in communication barriers (Grandpierre et al, 2018).

To sum up, the onion metaphor of culture is a representation of the layered nature of human culture, spanning from the visible through to the invisible cultures that shape interpretation and meaning making within the broader tapestry of quality healthcare delivery.

## Methodology

This section discusses the philosophical underpinnings, study approach, study design, sampling techniques, sample size, data collection method and analysis.

### Philosophical Underpinnings

This study is situated withing the anti-positivist paradigm. This philosophical viewpoint opposes the use of positivist methods in the social sciences to obtain reality (Wati, 2024). This suggests that anti-positivism focuses on qualifying rather than quantifying. Since the purpose of this study is to explore the lived experiences of patients and healthcare professionals regarding communication breakdowns in healthcare delivery in the Northern Region of Ghana, the researchers explored the deeper meanings and impacts of happenings during healthcare delivery. In light of this, Lim (2024) claims that because human behaviour is impacted by social surroundings, cultural settings, and subjective viewpoints, it cannot be fully studied using experimental perception and quantitative data alone. This line of thinking promotes subjective research methods above quantitative ones by highlighting the importance of comprehending the connections people make between their behaviors and relationships.

The northern Region of Ghana has unique sociocultural customs that influence the delivery of healthcare. Anti-positivism encourages analysts to consider these pertinent elements, which include cultural customs, traditional health beliefs, and community norms that might not be adequately represented by quantitative metrics alone (Iyadurai, 2023). The researchers were able to gather information on how these factors contribute to communication difficulties in hospital settings by using qualitative techniques like interviews.

### Study Approach

The qualitative research approach was adopted for this study. Hennink et al. (2020), averted that qualitative approach is a methodological technique that aims to comprehend human experiences and the interpretations that individuals make of things. In contrast to quantitative research, which analyses and assesses data through quantifiable analysis, qualitative research aims to probe the complexity of the problems by examining participant experiences (Hennink et al., 2020). This method works particularly well in medical settings where effective communication is essential to providing high-quality care. Because of this, the data is generated at the Tamale Teaching Hospital, Yendi Hospital, and Bimbilla Hospital respectively. In order to uncover the complex ways that communication impediments appear in actual circumstances the hospitals, interviews were conducted with patients and medical staff at Tamale Teaching Hospital, Yendi Hospital, and Bimbilla Hospital.

Through this qualitative study, the researchers were able to investigate the institutional and sociocultural framework in which those barriers are present. For example, the Tamale Teaching Hospital, Yendi Hospital, and Bimbilla Hospital settings allowed the researchers to understand some cultural nuances and how these created barriers to communication.

### Study Design **–** Phenomenology

The study design used in this investigation is phenomenology. This qualitative research method aims to comprehend and explain a person’s life experiences (Lindseth & Norberg, 2022; Williams, 2021). Shipp and Jansen (2021) highlight how people interpret their experiences and stress the subjective nature of reality. Phenomenology can offer important insights into how patients and healthcare providers engage in the context of health communication, particularly in a culturally diverse setting like Northern Ghana.

Health communication in the Northern Region of Ghana is influenced by distinct cultural, social, and economic elements (Abdulai, Ibrahim, & Anas, 2023). This suggests that culture is both visible and invisible, and that the person experiencing the occurrence has the best chance of interpreting it. In particular, this study required the use of this design because both patients and physicians provided narratives about their individual experiences. A phenomenological study design enables researchers to explore patients’ and healthcare workers lived experiences in greater detail (Walløe et al. (2024).

### Case Selection-Referral Hospitals in the Northern Region of Ghana

Northern Region of Ghana is geographically well suited for research of this nature due to the diverse cultures and multilingual communities located around the hospitals. By selecting the Tamale Teaching Hospital, Yendi Hospital and Bimbilla Hospital, this study covers a wide geographical area, incorporating both urban (Tamale) and rural (Yendi and Bimbilla) settings in the Northern Region of Ghana. The multicultural composition of patients and health care providers allows for a comprehensive understanding of effective communication in diverse settings. For instance, the Tamale Teaching Hospital serves as a referral center, treating many complex cases, while the Yendi and Bimbilla hospitals serve more local communities with unique cultural and communication challenges. This diversity provides valuable insights into how communication takes place between patients and healthcare providers and highlights its impact on the quality of healthcare delivery. Understanding how these dynamics influence patient-healthcare provider communication reveals important insights into the cultural barriers patients face in accessing health care. Therefore, the selection of Tamale Teaching Hospital, Yendi Hospital, and Bimbilla Hospital provide comprehensive approach to examining patient-healthcare provider communication in the Northern Region of Ghana due to their geographic, cultural and demographic diversity.

### Study Participants and Data Collection Procedures

The study population included a diverse group of participants, including patients and medical staff from the Tamale Teaching Hospital, Yendi Hospital, and Bimbilla Hospital. As previously said, the Tamale Teaching Hospital is the primary referral facility for the five Northern Ghanaian provinces, which makes it a suitable location to comprehend the complexities of health communication. Purposive sampling was used to pick healthcare providers to get a variety of participant experiences. In terms of the inclusion criteria, the study included patients who had recently sought medical attention at the selected hospital and as well as healthcare professionals who have lately worked at the facility.

After seeking clearance from the university and the hospitals, In-depth interviews were conducted with a varied sample of patients and providers at the Tamale Teaching Hospital, Yendi Hospital, and Bimbilla Hospital. The semi-structured nature of the interviews allowed for flexibility throughout the interactions, which ensured that pertinent information on communication difficulties was presented. The interviews examined participants’ interactions with members of the healthcare community, citing problems that hindered the open exchange of information. After meeting the 20^th^ participant, the researchers observed that no new trends emerged, only a repetition of narrations of previous participants. So, the researchers had to end the data collection process. The table below highlights the profile of the study participants

**Table 1.** Profile of Research Participants.

| S\N | Sex | Patients and Providers TTH-Tamale |
| --- | --- | --- |
| 1 | F | HPT1 |
| 2 | M | HPT2 |
| 3 | F | HPT3 |
| 4 | F | HPT4 |
| 5 | M | HPT5 |
| 6 | M | PPT1 |
| 7 | F | PPT2 |
| 8 | F | PPT3 |
| 9 | M | PPT4 |
| **Patients and Healthcare Providers-Yendi Hospital** |  |  |
| 1 | M | HPY1 |
| 2 | F | HPY2 |
| 3 | M | HPY3 |
| 4 | F | HPY4 |
| 5 | F | PPY1 |
| 6 | F | PPY2 |
| **Patients and Providers -Bimbilla Hospital** |  |  |
| 1 | M | HPB1 |
| 2 | F | HPB2 |
| 3 | M | PPB1 |
| 4 | F | PPB1 |
| 5 | F | PPB |

### Ethical Considerations and Informed Consent

Before the interviews, the researchers applied for introduction letter from the Faculty of Communication and Media Studies, UDS, Ghana. Also, ethical approval letter was granted by the ethical committee of the FCM, UDS, Ghana with ethical approval no: FCM\06\148. The introduction letter was attached to the ethical clearance letter for submission to the health facilities to facilitate entry into the field. The Tamale Teaching Hospital and the two other referral hospitals permitted us to collect the data. After this, the research participants were approached, and a frank and cordial meeting was scheduled for the time for the interview after they consented to participate. Before the commencement of the interviews, discussions were held on the purpose and rationale of the research to the study participants. Also, we sought their permission and abreast them of the interviews and recorded the process verbatim. The interviews were transcribed and utilized solely for academic purposes. Moreover, the interviews were conducted in a friendly and non-threatening environment and participants were allowed to have full control of the interview process in a manner that engendered trust and confidence. To protect the participants’ identities, represented each participant with a unique code, and no further details of their identification are given.

## Data Analysis-Themes identification

In this study, data analysis was conducted using both content and thematic analysis to derive meaningful insights from the qualitative data collected with patients and healthcare providers at the Tamale Teaching Hospital, Yendi Hospital and Bimbilla Hospital respectively. These two methods of qualitative data analysis complement each other and provide a comprehensive framework for understanding the effects communication barriers in healthcare settings. Riffe et al. (2023) state that content analysis is a carefully designed research method that is used to interpret textual data by noting and analyzing recurring and dominant words, themes or ideas within the data. Tracy (2024) recommends that content analysis allows researchers to identify patterns in communication, giving an organized way to analyze qualitative data.

The following steps were employed in the content analysis utilizing the Voyant Tools Software (VTS). The themes identification process began with importing the transcribed data into the VTS, specifically using the **Cirrus, Links, Trends** features. To identify major themes for the analysis, we extracted frequently used words by the participants using Voyant Tools Software (see Figure 1 and 2 below).

**Figure 1:**
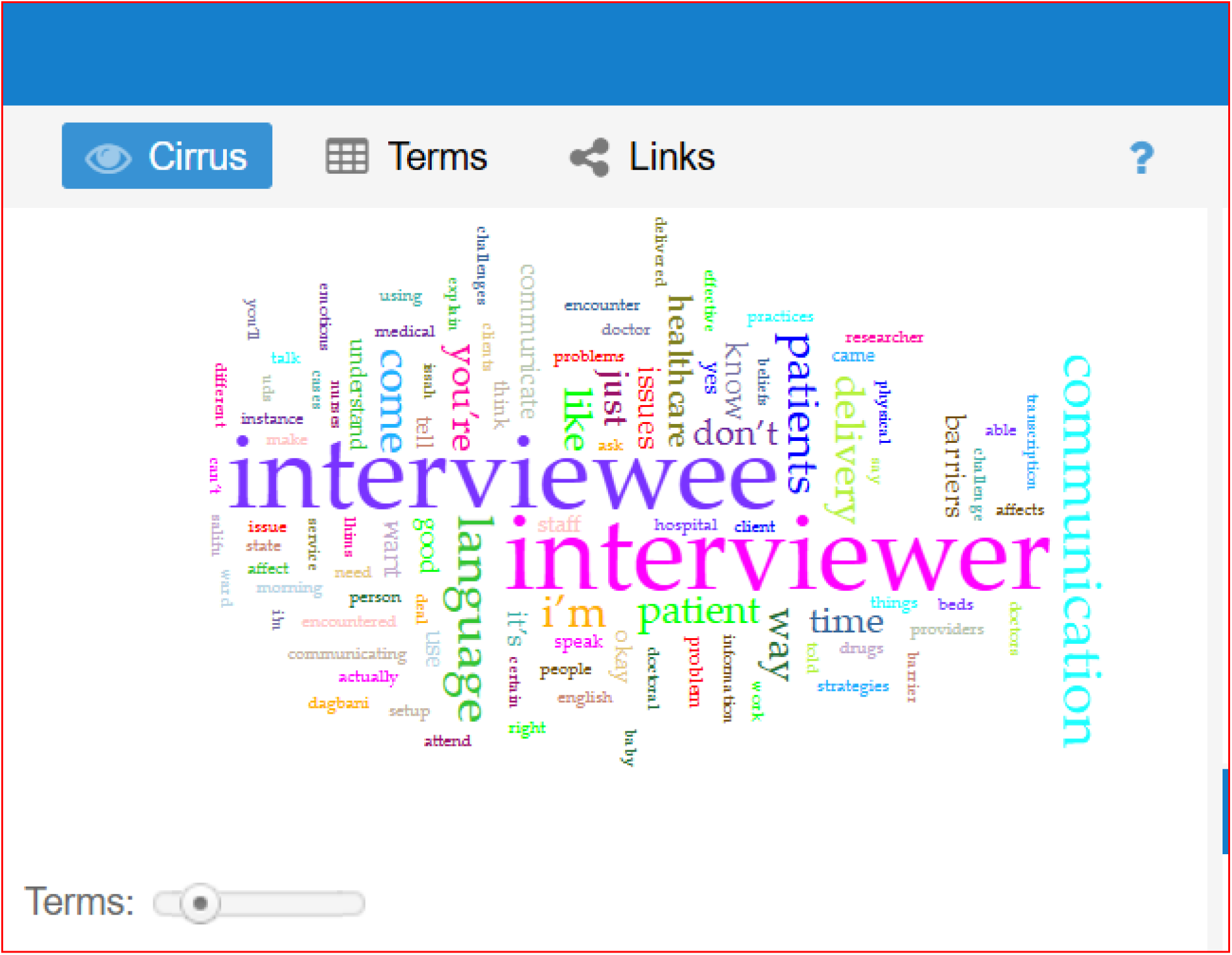
**Themes Identification utilizing the Cirrus**

**Figure 2:**
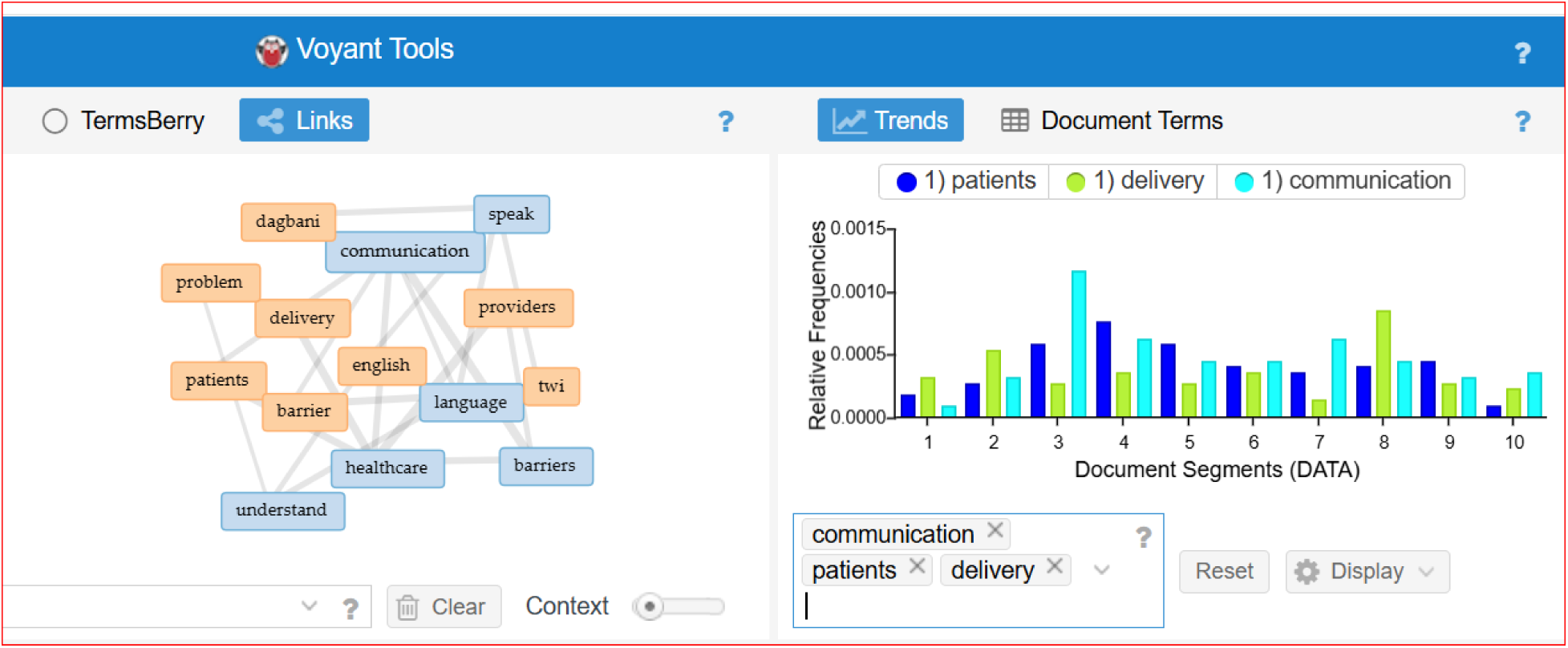
**Themes Identification utilising Links and Trends**

Figures 1 & 2 above display words frequently mentioned by the participants and their interconnections with other keywords in the data. The keywords captured in the Cirrus column include “Medical,” “Barrier,” “Errors,” “Time” and others. In the Links column, the highlighted keywords are “Language” “Communication”,” Speak” Understanding,” Problems”, “Healthcare” and the Trend Column-Patients, delivery, and Communication (Refer to Figures 1& 2 above). After carefully synthesizing the recurrent words, we constructed the following categories in the table below.

**Table 2.** Themes Construction.

|  |  |
| --- | --- |
| 1 | Misunderstandings and Medical Errors |
| 2 | Healthcare, Treatment and Increased Complications |
| 3 | Time, delivery, treatment |

*Source: Researchers` construct*

Given this, we manually combined the frequently mentioned words and the constructed categories and manually formed the main themes for the analysis. Thus, frequently Used Words + Constructed Categories =Main Themes.

Theme1: Misunderstandings and Misdiagnoses: A Layered Communication Challenge

Theme 2: Delayed Treatment and Increased Complications: Systemic Failures Beneath the Surface

Theme 3: Poor Adherence to Treatment Plans

## Findings and Discussion

This study examines how communication breakdowns, particularly within Ghanaian clinical environments, shape both the experiences of patients and the outcomes of care. Drawing on perspectives from healthcare providers and service users, it reveals how language barriers, cultural misunderstandings, and systemic inefficiencies interfere with effective treatment. The dominant themes explored include misdiagnoses, delayed treatment, reduced patient satisfaction, and poor adherence to medical instructions.

### Misunderstandings and Misdiagnoses: A Layered Communication Challenge

The theme of misunderstandings and misdiagnoses highlights the profound impact of communication breakdowns in clinical encounters. For instance, one of the participants said.,

*“…sometimes instead of examining up to 10 patients, you just feel reaching 4 is enough, after all the communication stress is too much.We are so tired that when you want to even close at work patients are still waiting. Because of this we are sometimes not able to give all attention (HPT1).”*

This captures how ongoing communicative strain can demotivate healthcare providers, leading to reduced diligence and incomplete care routines. This quiet disengagement is often not a sign of professional negligence but rather a symptom of deeper frustrations rooted in unresolved cultural, emotional, and linguistic tensions (Gerard, 2024).The Onion Metaphor, an intercultural communication model used to conceptualize human interactions in layers, offers a helpful lens for unpacking the various dimensions of miscommunication (Aleksandrova et al., 2024). The outermost layer includes observable behaviors such as shortened conversations, fragmented instructions, and patient withdrawal. The intermediate layer encompasses healthcare norms, provider assumptions, and patient expectations shaped by societal and institutional influences. At the core, however, lie unspoken patient fears, provider burnout, and deep-seated cultural or religious beliefs. These are the forces that silently influence clinical interactions.

This is clearly demonstrated in HPY1’s account:

> *“For instance, in giving medications we are expected to explain to the patient why that medication must be given, but for language barrier we just give the medications without telling the patient why they have to take that medication. Other times too, patients want to ask why they are giving certain medications but how to communicate their concerns are a problem because you both don’t understand each other’s language.*

Here, a surface-level language barrier masks a more serious issue, which is the erosion of patient agency due to an inability to meaningfully engage with the provider (Gundersen and Bærøe, 2022). Such miscommunications hinder participatory care and raise the risk of diagnostic and treatment errors. Supporting evidence from similar contexts reinforces these observations. Studies in Ghana and East Africa consistently show that language barriers and the use of untrained interpreters often result in shorter consultations and clinical misjudgments (Peterson, 2023; Mwasaru, 2022). Yet, beneath these logistical issues lies a human story. Patients rehearse their responses, select providers based on perceived empathy, and conceal symptoms. All of these are coping mechanisms shaped by fear, mistrust, and historical power imbalances. As HPY1 explains, *“…patients communicate among themselves and have rehearsed answers…They study and select those they think are calm.”*

This kind of strategic concealment serves as a defense mechanism that preserves patients’ dignity in culturally alienating environments but distorts the medical dialogue. Likewise, HPB1, notes, *“…the wrong use of words by interpreters and even the context might differ from the intended purpose of the discussion,” emphasizes* how subtle semantic mismatches could lead to significant diagnostic consequences.

The onion metaphor reminds us that what seems like a simple issue, such as an abbreviated consultation, an incomplete medical history, or a delayed diagnosis, is merely the surface. Addressing such problems requires a willingness to peel back the layers through culturally competent and patient-centered communication that treats the person, not just the condition.

### Delayed Treatment and Increased Complications: Systemic Failures Beneath the Surface

The theme of delayed treatment and increased complications illustrates how communication barriers, cultural practices, and logistical challenges intersect.

HPT4 account,

> *“A lady once came here with a condition and was referred to the Korlebu Teaching hospital, but she declined and said she had to go home and do consultations after the referral letter was prepared. She later returned to accept the referral, but her condition was worsened at the time she returned, to a point her condition couldn’t be managed any longer.”*

This demonstrates how cultural obligations and the need for family consultation can override clinical urgency. While this decision may seem irrational from a biomedical perspective, it becomes understandable when viewed through the deeper layers of the onion metaphor, which are shaped by cultural scripts, gender roles, and collective decision-making. At the middle layer, healthcare communication often fails to convey urgency or risk effectively, especially when language barriers exist. Patients may receive referral letters or discharge instructions without fully understanding their implications. This often results in delayed decisions and worsened clinical outcomes. These failures are made worse by infrastructural shortcomings and overburdened staff who may lack the time or tools to explain medical urgency in culturally meaningful ways.

Caregivers are also heavily burdened as they navigate the healthcare system with limited support. PPB1’s experience,

> *“ The first challenge with that has to do with if you’re the only relative to the sick patient. Abandoning the patient and leaving the hospital to go and purchase the drug, your mind will not be at ease, especially if the patient’s condition is critical.”*

reflects the emotional and logistical strain placed on caregivers. The requirement to leave a patient’s bedside to obtain basic supplies reveals a deeper issue of institutional disengagement. These logistical shortcomings may occupy the outermost layer of the onion metaphor but carry significant emotional and ethical implications rooted in structural neglect.

HPT5’s report of supply shortages,

> *“…Because sometimes you can come to work and there are no hand gloves yet you’re expected to set up something for a client, at that moment you’re compelled to use bare hands. You can even use your bare hand and when you’re done there isn’t water to wash your hands.”*

further highlights the tension between professional ethics and material constraints. These shortages not only disrupt clinical procedures but also damage staff morale, leading to corner-cutting and delayed interventions that ultimately endanger patient safety.

Perhaps the most severe example comes from HPY4, who recounted,

> *“I was here when they brought somebody with a minor cut that required operation, it took them a week to attend to the person…the leg got badly infected that we had to cut it off. Something should have been done faster to help that person but here, it’s not like that. How to even go for blood test and other things delay too much”*

This case reflects a series of failures including poor communication of urgency, administrative delays, lack of supplies, and potentially flawed triage processes. More deeply, it reveals a healthcare system in which patient suffering is normalized and individual pain is depersonalized.

### Poor Adherence to Treatment Plans

The theme of poor adherence to treatment illustrates how cultural beliefs, ineffective communication, and lack of patient education combine to influence patient behavior and treatment outcomes. These insights from frontline healthcare providers reveal how communication failures and social factors pose serious challenges to adherence in Ghana’s healthcare system.

One major factor is the influence of cultural and religious beliefs on patients’ willingness to follow medical advice. HPT2 explains,

> *“…some patients refuse certain medical practices due to their faith or beliefs. An example is some refusing blood transfusions because their religion is against it. Someone came here for treatment and was anemic, when the doctor requested blood transfusion, she said her religion does permit that one.”*

This is consistent with Mensah (2021), who found that some Pentecostal and charismatic groups in Ghana reject biomedical interventions based on religious teachings. These beliefs can seriously hinder medical reasoning and outcomes if not properly addressed with sensitivity.

Another barrier is the mismatch between patient expectations and the realities of medical care. HPT2 notes,

> *“…some clients expect magic when you prescribe drugs for them. You need to let them understand how the process works so they don’t assume different things. Many of them simply do not understand how the treatment plans are carried out and sometimes, I understand them because of the pain they are going through”*

This highlights the need for better patients’ education. Viswanath (2024), in a study on Indian mothers and trauma, found that many patients misunderstood prescriptions and expected instant recovery, often stopping medication early. Similarly, Mathebula (2023) found in South Africa that poor explanations about dosage and outcomes led to high non-compliance, especially in chronic care. Unrealistic expectations, when unaddressed, can weaken the therapeutic bond between patients and providers.

Refusal to follow treatment, even when unrelated to religion, is also a concern. HPB2 recounts,

> *“…a patient can be on a sick bed and refuse taking medications anytime you want to administer drugs. It could sometimes be due to the fact that they are allergic to certain drugs from their past experiences, but they can’t explain that to you.”*

This shows the challenge of interpreting silent resistance, which may result from previous negative medical experiences. Chakaya et al. (2021) found similar issues where patients failed to disclose past drug reactions, leading to treatment refusal often misinterpreted as stubbornness.

The human aspect of communication barriers is especially visible in gender-sensitive situations. HPY3 observes,

> *“…if the person isn’t opening up and just resisting, that is where we have a challenge…some ladies would tell you because of this and that reason I don’t want you to examine me this way. Others will not say anything against your directive”*

This reflects a deeper discomfort linked to gender norms and personal boundaries. In Ghana, Ampofo and Boateng (2020) observed that gender relations shape health interactions, with many women resisting physical exams due to fear or trauma. Similarly, Onyango et al. (2017) found in Kenya that women avoided certain medical examination because of modesty and discomfort with male providers. These concerns are rarely addressed with patient-centered communication leading to worsening disconnect.

## Summary of key findings

According to the study, communication obstacles in healthcare are more than just minor issues; they are deeply layered challenges with far-reaching consequences. Misdiagnoses and insufficient clinical assessments are frequently the result of patient-provider misunderstandings. These visible flaws, however, are indications of deeper concerns rooted in cultural disconnects, emotional strain, and institutional exhaustion.

One major conclusion is how language barriers and reliance on informal interpreters jeopardize diagnosis accuracy and undermine confidence. Patients who are unable to adequately express their problems may hide essential information or engage in prepared encounters to control perceived power imbalances. Such actions distort therapeutic conversation and impede participation in care.

Delayed treatment appears as another key worry, frequently caused by miscommunication of urgency, cultural consultation norms, and logistical challenges. Care providers experience mental and physical hardship when navigating fragmented systems, sometimes leaving patients unattended to obtain essential necessities. These delays are exacerbated by material shortages and infrastructural neglect, which reduce clinician responsiveness.

The study also emphasizes the emotional impact of contemptuous communication. Patients report feeling ignored or intimidated by provider attitudes, which leads to dissatisfaction and a breakdown in the therapeutic relationship. This discontent is increased when physicians fail to communicate clearly, empathically, or timely, particularly during important moments of diagnosis or referral.

Finally, a lack of culturally relevant health information, as well as prior unpleasant experiences, frequently disrupt treatment adherence. Religious convictions, unmet expectations, and unresolved previous traumas frequently cause silent resistance or open refusal of care. These issues are especially severe in gendered settings, as modesty and fear prevent free communication during physical examinations.

Overall, the data show that effective healthcare communication requires more than just linguistic translation. It necessitates cultural and emotional intelligence, as well as systemic improvements that acknowledge patients as individuals immersed in complicated social and psychological realities, rather than cases to be controlled.

## Conclusion

The study concluded that communication obstacles have a significant impact on patient outcomes and satisfaction in Northern Ghana’s healthcare settings. The study established that misconceptions and misdiagnoses are not isolated clinical blunders, but rather the result of ongoing linguistic and cultural gaps between healthcare providers and patients. These communication breakdowns jeopardized proper diagnosis, lowered the quality of care, and decreased patient trust in the healthcare system. The study also revealed how healthcare providers who are overwhelmed by the strain of inefficient communication withdraw from full clinical engagement. This silent resignation, evident in incomplete procedures and limited empathy, indicates systemic strain rather than individual wrongdoing. Patients, in turn, employ coping techniques such as rehearsed responses, selective disclosure, or utter silence, all of which jeopardized the accuracy of clinical assessments and the quality of care provided. Furthermore, relying on informal and inexperienced interpreters adds complexity, frequently distorting medical communication and resulting in poor therapeutic outcomes.

## Recommendations

It is recommended that the Ghana Health Service (GHS) work with regional health directorates and professional health training institutions to incorporate comprehensive communication and cultural competence training. The training should reflect in both pre-service curricula and in-service capacity-building programs for healthcare providers. This training should cover not only verbal clarity and empathy, but also recognizing and respecting cultural values. By institutionalizing such training, GHS could provide healthcare personnel with the skills they need to develop trust, improve treatment adherence, and increase overall patient happiness in a variety of clinical settings.

Also, GHS should recruit professional interpreters with backgrounds in science communication and intercultural communication competence to facilitate the free flow of communication in language barrier situations.

Again, more trained providers should be deployed to the facilities to make up for the deficit in patient-provider ratio that often led to provider burnout that culminates in rushed diagnosis and poor attitudes towards patients.

## Direction for Further Studies

To conduct further studies, the following suggestions are outlined: the Again, the study should examine code mixing and code switching between patients and healthcare providers at the referral hospitals in the Northern Regions of Ghana. Lastly further research should explore strategies for effective communication at the referral hospitals in the Northern Region of Ghana.

## Ethical Approval

All procedures performed in this study were in accordance with the ethical standards of the University for Development Studies, Ghana. The name of the ethical committee is the Ethical Review Board of University for Development Studies, Ghana.

## Informed Consent

was sought from the management of the hospitals because human participants were involved.

## Authors’ Contributions

The first author conceptualized and designed the study and collected the data and drafted the article and supported in analyzing the data. The second author also supported in analyzing the data and made some suggestions about theories and the methodology of the manuscript. The third author reviewed the literature and edited the manuscript. All authors reviewed and approved the final version of the manuscript.

## Data Availability

The data supporting the findings of this study are available from the corresponding author, Salifu Issah, upon reasonable request.

## Data Availability

Data for this study were from interviews and they have been transcribed.

